# Hypothesis-Driven Thalamic Stereo-EEG Informs Surgical Therapy for Drug-Resistant Epilepsy: Outcomes in 117 Consecutive Patients

**DOI:** 10.64898/2026.11.23.26368733

**Authors:** Suk Joon Lee, Brian Hsueh, Lidia Moura, Catherine Chu, Peter Hadar, Alex Bender, Lily Grossmann, Elizabeth Thiele, Lara Basovic, Alice Lam, Andrew J. Cole, Sydney Cash, Pranav Nanda, R. Mark Richardson

**Author notes:** S.L. and B.H. contributed equally to this work.

## Abstract

**Objective:** Thalamic stereoelectroencephalography (SEEG) is used to characterize seizure networks and guide neuromodulation, yet its safety and clinical utility are incompletely characterized. We examined how thalamic SEEG informs surgical therapy.

**Methods:** We reviewed 117 consecutive patients who underwent thalamic SEEG at Massachusetts General Hospital (2020–2025), in whom the anterior (ANT), centromedian (CM), or pulvinar (PUL) nucleus was sampled to test a seizure network hypothesis. We assessed complications, targeting accuracy, the effect of thalamic ictal timing on laterality determination and neuromodulation target selection, and seizure outcomes.

**Results:** Small hemorrhages were seen on postoperative imaging for 0.3% of leads (4/1507); all were asymptomatic and none involved a thalamic lead. Targeting accuracy for ANT, CM, and PUL was 81.0%, 94.2%, and 100%, respectively. Early bilateral thalamic ictal signals identified bilateral seizure networks with 75.6% sensitivity and 78.1% specificity, rising to 92.6% sensitivity when bitemporal onset was excluded. Among 52 patients planned for thalamic or thalamocortical responsive neurostimulation (RNS), the pre-SEEG target was revised in 34.6%, including in patients whose cortical hypothesis was unchanged. Most cortical seizure-onset zones did not map to a single thalamic nucleus. Among patients treated with thalamic or thalamocortical RNS, those with early thalamic ictal signals had better seizure outcomes than those without (standardized mean difference 1.71, 95% CI 0.77-2.65). Mean primary seizure reduction was 75.0 ± 31.7% after RNS (seizure freedom 40.4%) and 90.9 ± 20.6% after resection or laser interstitial thermal therapy (61.8%).

**Significance:** Thalamic SEEG has a safety profile similar to that of non-thalamic SEEG, provides patient-specific information that informs neuromodulation strategy and target selection, and identifies an early thalamic ictal signal that is associated with better seizure outcomes. These findings support hypothesis-driven thalamic sampling in the evaluation of drug-resistant epilepsy.

**KEY POINTS:**

- No hemorrhage occurred with 373 thalamic SEEG leads, and ANT, CM, and PUL were targeted accurately.
- Early bilateral thalamic ictal signals identify bilateral seizure networks (75.6% sensitivity, 78.1% specificity).
- The thalamic RNS target changed in 34.6% of RNS candidates, including in patients whose cortical hypothesis was unchanged.
- Cortical seizure-onset zones do not map exclusively to a single thalamic nucleus.
- An early thalamic ictal signal on SEEG is associated with better outcomes after thalamic RNS.

## INTRODUCTION

As a central relay hub integrating widespread cortical and subcortical networks, the thalamus is an important target for neuromodulation in patients with drug-resistant epilepsy.^1^ Multiple studies have demonstrated that modulation of specific thalamic nuclei, including the anterior nucleus (ANT), ^2,3^ centromedian nucleus (CM), ^4–6^ and pulvinar nucleus (PUL) ^7,8^, can lead to meaningful improvements in seizure outcomes. Given the efficacy of thalamic neuromodulation, stereoelectroencephalography (SEEG) is increasingly used to characterize thalamic involvement in patient-specific seizure networks, ^9^ with findings that inform both resective surgery planning and chronic neuromodulation strategies.^10^

Registry data show the clinical importance of patient-specific thalamic targeting. In the Medtronic Registry for Epilepsy (MORE), ANT deep brain stimulation (ANT-DBS) produced a median seizure frequency reduction of only 33% at two years when applied broadly across epilepsy types, compared with 56% in the SANTÉ pivotal trial.^11^ This discrepancy was attributed in part to the lower proportion of temporal lobe epilepsy patients in MORE (39% versus 60% in SANTÉ), whose seizures more often propagate through the Papez circuit that ANT modulates. Even within temporal lobe epilepsy, thalamic recruitment is not uniform. A recent study of multisite thalamic SEEG recordings reported that while ANT was engaged first in most hippocampal-onset seizures, the PUL led in the majority of non-hippocampal-onset seizures, so structural connectivity alone does not predict which nucleus is the most effective neuromodulation target.^12^

At our center, responsive neurostimulation (RNS) is the default neuromodulation strategy for patients whose seizure network cannot be resected or ablated. Because detection and stimulation are programmed per lead, the contacts that see the seizure first can trigger therapy anywhere in the implanted network, so the SEEG evaluation must answer three questions: where epileptiform activity should be detected to trigger stimulation, where stimulation should be delivered, and whether and when the thalamus becomes involved.^13^ Thalamic SEEG at our center evolved from this need, guided by the principle that the latency of thalamic recruitment relative to cortical onset determines the RNS configuration. Where thalamic involvement is delayed, the sampled nucleus is unlikely to afford early detection, and a corticocortical configuration may be favored for neuromodulation. Where there is little or no lag, the thalamus likely participates in the primary organization of the seizure rather than receiving propagated activity, and a thalamocortical or bithalamic configuration is indicated, according to whether that organization is unilateral or bilateral. Recruitment timing therefore converts an anatomical hypothesis into a patient-specific comparison among the therapeutic configurations, permitting selection of the one most likely to disrupt each patient’s seizure organization. Despite this rationale, the literature on thalamic SEEG consists largely of small case series. One relatively large cohort study (74 patients) demonstrated thalamic involvement in seizure networks in 86% of patients, but the impact on subsequent treatment decision-making and the safety profile of thalamic SEEG implantation were not systematically evaluated.^14^ Both the safety and the clinical utility of thalamic SEEG therefore remain incompletely defined.

We analyzed 117 consecutive patients who underwent thalamic SEEG at our institution. We evaluated safety, targeting accuracy, and seizure outcome, and asked whether thalamic SEEG findings change the laterality hypothesis and the neuromodulation target, and whether the timing of thalamic recruitment is associated with seizure outcome.

## METHODS

### Data selection

This was a retrospective cohort study of consecutive patients. We reviewed SEEG targeting records for all patients who underwent SEEG implantation at Massachusetts General Hospital between January 1, 2020, and December 31, 2025. Among these patients, 117 had at least one SEEG depth electrode (lead) implanted in the thalamus and were included. The study was approved by the Mass General Brigham Institutional Review Board, which waived the requirement for informed consent for this retrospective review.

All 117 patients were included in the analyses of safety and thalamic lead targeting. Clinical and demographic information was extracted from the electronic medical record. Of these, 108 patients had spontaneous seizures captured during SEEG and were included in analyses of seizure-onset hypotheses, seizure laterality, and post-SEEG treatment decision-making. Of these, 103 underwent therapeutic surgery and 86 had seizure outcome assessment at least 6 months after therapeutic surgery.

### Postoperative complications and thalamic SEEG targeting analysis

Postoperative hemorrhagic complications were identified from radiology reports of post-implantation MRI or CT.

Preoperative and postoperative imaging were processed using Lead-DBS^15^ and 3D Slicer^16^ software. The coordinates of the centroid of each SEEG contact were calculated in Montreal Neurological Institute (MNI) space. Each contact was assigned to the THOMAS atlas nucleus within a 2-mm radius of its centroid, allowing for reconstruction and normalization uncertainty.^17^ For 12 patients in whom spatial normalization failed, preoperative MP2RAGE MRI and postoperative CT were coregistered in Visage Imaging and the overlap between contacts and the target nucleus was assessed visually.

### Seizure etiology and hypothesis review

Epilepsy etiology was classified hierarchically as *MRI-lesional* (SEEG-defined seizure-onset zone concordant with a structural lesion), *genetic* (confirmed pathogenic diagnosis), *post-infectious*, *post-traumatic*, or *post-hemorrhagic* (documented antecedent event), or *unknown*.

Seizure type was determined from the post-SEEG multidisciplinary conference discussion, seizure history, electrographic findings, and semiology according to the most recent International League Against Epilepsy (ILAE) classification.^18^ Epilepsy was classified as focal, generalized, or unknown. Focal epilepsy was further subdivided into *local*, *regional*, and *multiregional* seizure-onset patterns. *Local* was defined as seizure onset confined to a single lobe; *regional* as broad seizure onset spanning contiguous lobes; and *multiregional* as multiple independent seizure-onset zones involving more than one lobe.

For pre-SEEG hypotheses, the hypotheses for (1) the cortical seizure-onset zone and (2) the thalamic nucleus first involved in seizure propagation were extracted from the multidisciplinary epilepsy surgery conference note before SEEG. When the note did not record them explicitly, the hypotheses were generated from the clinical data in the note, including semiology, imaging, phase 1 data, and neuropsychological assessment. This review was performed blinded to patient demographic information and subsequent therapeutic intervention.

### Assessment of thalamic ictal involvement

SEEG recordings from the epilepsy monitoring unit were visually reviewed using Natus software to determine the timing of thalamic nucleus involvement during ictal episodes. For patients with multiple seizure types, up to two were reviewed: one from each side for bilateral independent onsets, otherwise the two most clinically severe, prioritizing generalized tonic-clonic activity or loss of consciousness.

Thalamic involvement was defined as (1) early if the delay between cortical onset and thalamic nucleus ictal signal was less than one second, (2) late if the delay was one second or more, or (3) not involved if no clear visual electrographic signature within the electrographic ictal episode (between seizure onset and offset) was identified. For patients with SEEG leads targeting multiple thalamic nuclei, the nucleus demonstrating the earliest ictal signal (low voltage fast activity, rhythmic spikes, or rhythmic spike and wave discharges) was selected for analysis. When the relative temporal involvement of two or more nuclei could not be differentiated, the nucleus with the highest-amplitude signal was selected. The contact numbers corresponding to the thalamic contacts demonstrating the strongest signal were recorded for subsequent analysis. Display sensitivity for visual review of thalamic signal was 5–20 µV/mm.

Visual review of SEEG was not blinded to patient information.

### Therapeutic intervention and seizure outcomes

Operative reports were reviewed to determine the type and anatomical targets of subsequent therapeutic surgical interventions.

Baseline seizure frequency was recorded from the pre-SEEG epilepsy conference note or, when absent, from the neurology clinic note closest to the SEEG procedure. Postoperative seizure outcomes were assessed using the most recent neurology clinic note that recorded the frequency of the patient’s primary seizure type. The primary seizure type was defined as the most disabling seizure in terms of severity (generalized tonic-clonic seizure > focal seizure with impaired consciousness > focal seizure with preserved consciousness). Primary seizure reduction was defined as (baseline frequency − current frequency)/baseline frequency, expressed as a percentage.

### Statistical analysis

Continuous variables are reported as mean ± SD. Sensitivity, specificity, and predictive values are reported with Wilson score 95% confidence intervals. Seizure outcomes between groups were compared by the standardized mean difference, calculated as Hedges’ g: the difference in means divided by the pooled SD, multiplied by the small-sample correction factor J = 1 − 3/(4[n1 + n2] − 9). Its 95% CI used the normal approximation with SE = √[(n1 + n2)/(n1n2) + g²/(2[n1 + n2])]. A CI that excluded 0 was considered evidence of a between-group difference. All analyses were performed in Python.

## RESULTS

### Demographics

Among the 117 patients included in the study, 53 (45.3%) were female and 64 (54.7%) were male. The mean age at the time of SEEG implantation was 27.8 ± 14.8 (mean ± SD) years and the mean duration of epilepsy was 13.2 ± 9.3 years. Thirty-two patients (27.4%) had undergone at least one prior epilepsy surgery for drug-resistant seizures before thalamic SEEG implantation. Etiologies and SEEG-confirmed seizure-onset locations are summarized in Table S1.

### Evolution of hypothesis-driven thalamic SEEG strategies

The mean epilepsy monitoring unit (EMU) stay was 9.9 ± 4.6 days. A total of 1,507 SEEG leads were implanted in 117 patients, corresponding to a mean of 12.9 ± 3.5 leads per patient. Of these, 373 leads targeted the thalamus, including ANT, CM, PUL, or a combination, according to the pre-SEEG seizure network hypothesis (Figure 1A). Over six years, thalamic sampling broadened from CM alone to hypotheses that also sampled ANT and PUL (Figure 1B). Since the close of this cohort, the protocol has expanded further to include the mediodorsal nucleus, which adds non-overlapping frontal connectivity (Figure 1A).

**Figure 1.**
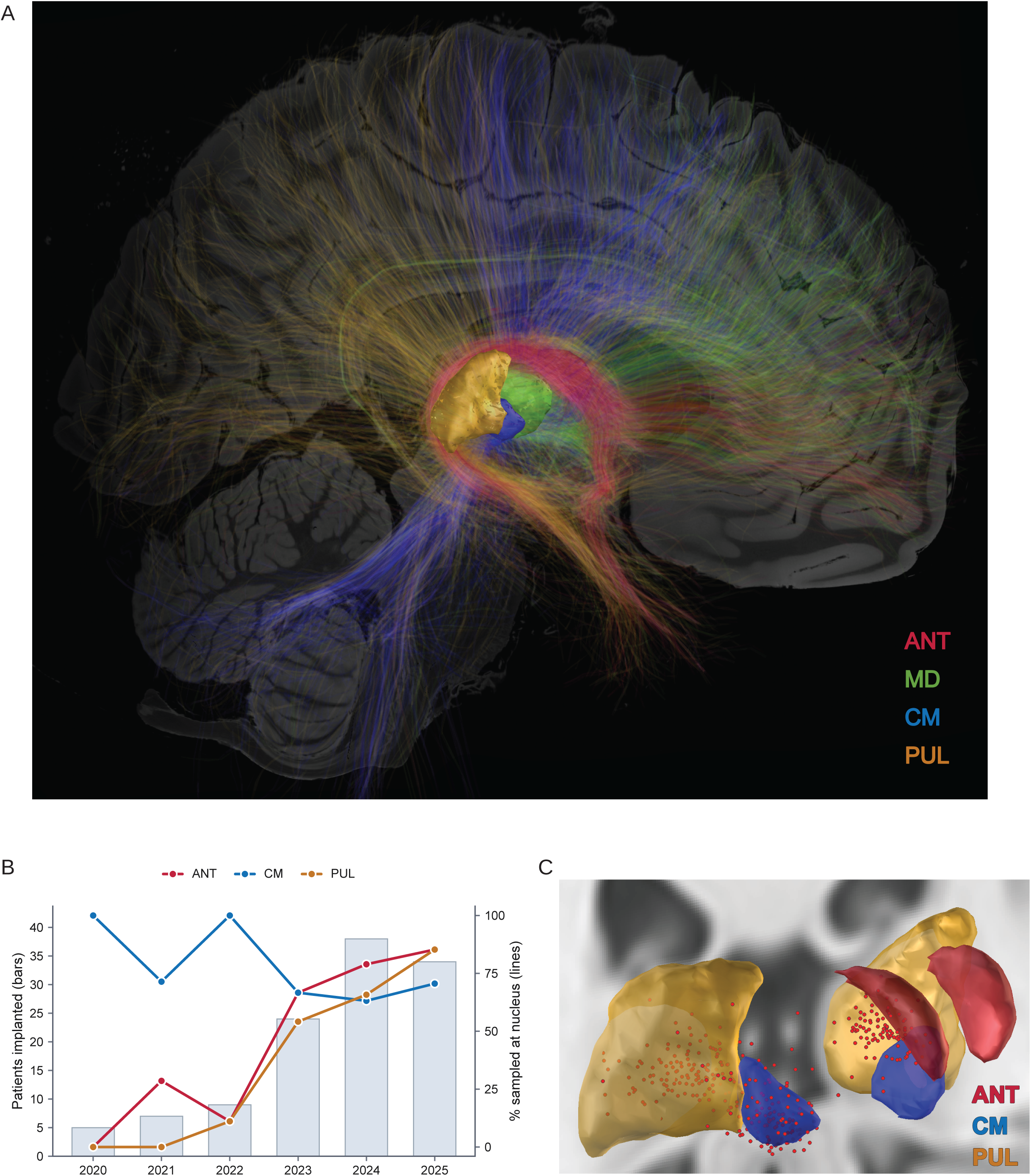
Thalamic sampling during SEEG: targets, growth, and lead placement. **(A)** Structural connectivity of the canonical thalamic targets on a Human Connectome Project template based on diffusion tractography. Each nucleus segmentation and its streamlines are shown in the same color: anterior nucleus (ANT, red), centromedian nucleus (CM, blue) and pulvinar (PUL, amber). The distinct cortical projection fields of each nucleus provide the anatomical rationale for selecting a thalamic target according to the hypothesized cortical seizure network. The mediodorsal nucleus (MD, green) was not implanted during this cohort but is now included in our thalamic SEEG protocol. **(B)** Thalamic SEEG implantation by year (n = 117 patients, 2020–2025). Bars give the number of patients implanted each year (5, 7, 9, 24, 38 and 34 for 2020 through 2025). Lines and circular markers give the proportion of that year’s patients in whom each nucleus was sampled (ANT red, CM blue, PUL amber). **(C)** Representative post-implantation reconstruction showing the contacts of thalamus-targeting SEEG leads that showed the strongest thalamic ictal signals on visual review of the SEEG recording (n = 170 leads with strongest-signal contacts). Left hemisphere leads were transformed to the right hemisphere coordinates for visualization.

### Safety and targeting accuracy of thalamic SEEG implantation

The overall hemorrhage rate was 0.3% (4/1507 leads), corresponding to 3.4% of patients (4/117). No complications occurred with 373 thalamic SEEG leads. Four intracranial hemorrhages associated with non-thalamic SEEG leads were identified on postoperative imaging, including three epidural hematomas measuring 4, 7, and 14 mm in maximum thickness and one subdural hematoma measuring 3 mm. All hemorrhages were asymptomatic and were managed conservatively without surgical evacuation.

Targeting accuracy was defined as the presence of at least one contact within the intended thalamic nucleus. Overall, targeting accuracy was 81.0% for ANT leads (102/126), 94.2% for CM leads (129/137), and 100% for PUL leads (110/110). Mean radial error from the planned trajectory for contacts outside the target nucleus was 1.31 ± 0.46 mm for ANT and 1.42 ± 1.35 mm for CM, within the expected range of SEEG implantation targeting accuracy.^19^

Among leads with at least one contact in the intended nucleus, we next asked whether the contacts with the strongest ictal signal lay within that nucleus. In every lead, at least one strongest-signal contact lay within the intended nucleus (ANT 46/46; CM 65/65; PUL 59/59), and in many leads (ANT 33/46, CM 56/65, PUL 13/59) additional contacts with equally strong signals lay outside it (Figure 1C).

### Thalamic SEEG assists laterality determination

Seventy-three patients underwent bilateral thalamic SEEG implantation. In 46 of these, the pre-SEEG hypothesis was unilateral, and SEEG revised laterality to bilateral in 15 (32.6%). Bilateral networks were defined as having (1) bilateral simultaneous cortical initiation or (2) independent seizure onsets originating in both hemispheres. Of these 15 patients, nine demonstrated early thalamic ictal signals bilaterally (Figure 2A). Five of the remaining six, whose laterality was revised without early bilateral thalamic signals, had bitemporal seizure onset.

**Figure 2.**
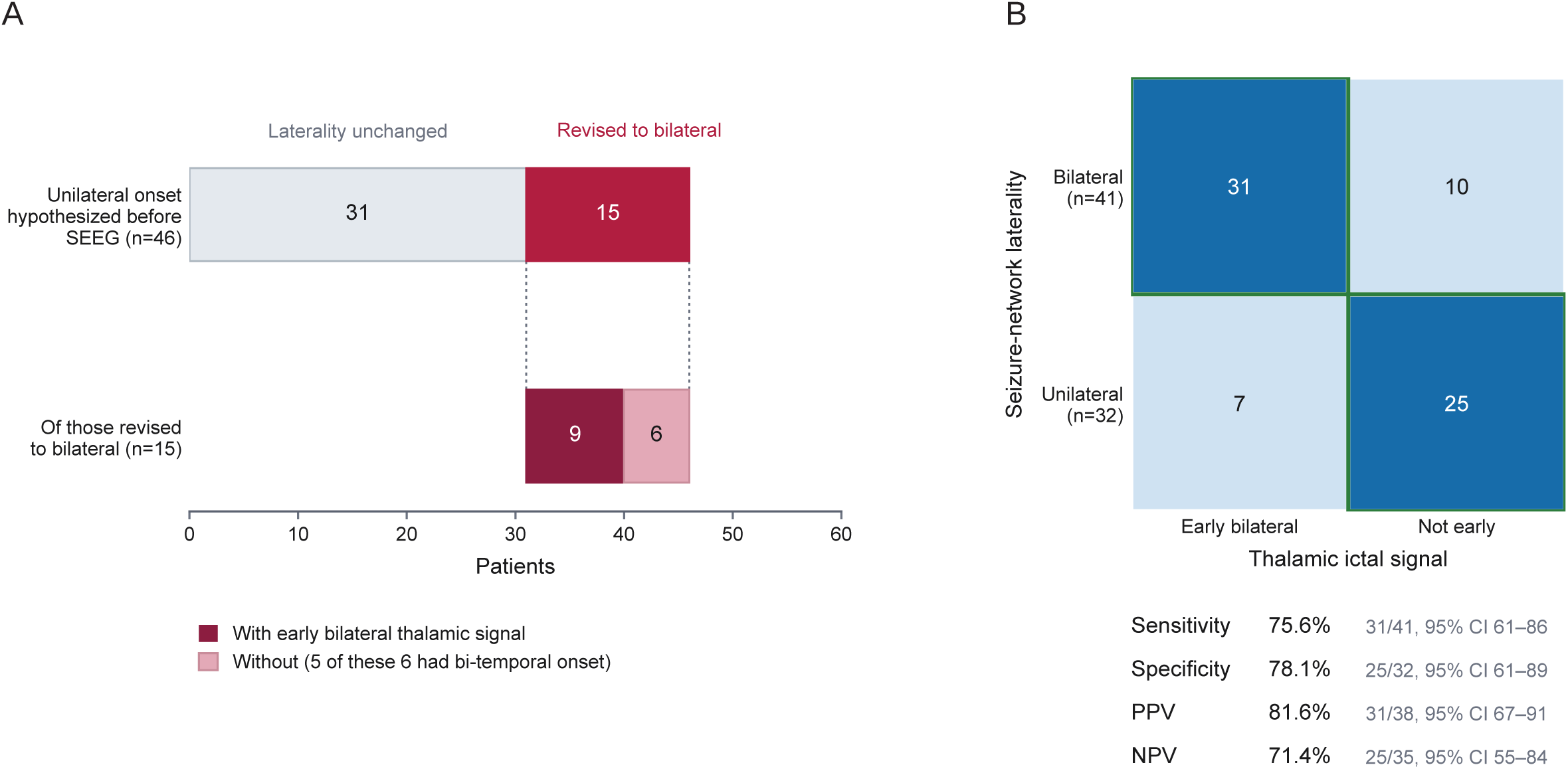
Early bilateral thalamic engagement identifies a bilateral seizure network. **(A)** Laterality outcome of pre-SEEG unilateral hypothesis cases (n = 46) where SEEG leads were implanted in bilateral thalami. The hypothesis was retained in 31 patients (grey) and revised to bilateral in 15 (red). The revised bar is subdivided by whether an early bilateral thalamic signal was present: nine of the 15 revisions were accompanied by an early bilateral thalamic signal, and six were not; five of those six had bitemporal onset. **(B)** Agreement between the thalamic ictal signal (early bilateral vs not early) and the SEEG-confirmed seizure-network laterality (bilateral vs unilateral) among all patients with bilateral thalamic implantation (n = 73). Of 41 patients with a bilateral seizure network, 31 showed an early bilateral thalamic signal and 10 did not; of 32 with a unilateral network, 25 showed no early bilateral signal and seven did. Taking the SEEG-confirmed seizure network laterality as the reference standard, the early bilateral thalamic signal had a sensitivity of 75.6% (31/41; 95% CI 60.7–86.2), specificity 78.1% (25/32 patients with SEEG-confirmed unilateral networks; 61.2–89.0), positive predictive value 81.6% (31/38; 66.6–90.8) and negative predictive value 71.4% (25/35; 54.9–83.7). In the post-hoc analysis excluding the 14 patients with bitemporal seizure onset (n = 59), sensitivity was 92.6% (25/27; 76.6–97.9) and specificity 78.1% (25/32; 61.2–89.0). Confidence intervals are Wilson score intervals.

We further evaluated the relationship between SEEG-confirmed seizure laterality and thalamic ictal signals among all 73 patients who underwent bilateral thalamic SEEG implantation. Early bilateral thalamic ictal signals identified bilateral seizure networks with a sensitivity of 75.6% (31/41, 95% CI 60.7-86.2) and specificity of 78.1% (25/32, 95% CI 61.2-89.0) (Figure 2B). Because bitemporal epilepsy did not consistently show early thalamic involvement (Figure 2A), a post hoc analysis excluding the 14 bitemporal patients gave a sensitivity of 92.6% (25/27, 95% CI 76.6-97.9) with unchanged specificity.

### Thalamic SEEG informs neuromodulation strategy and target selection

Among the 108 patients in whom spontaneous seizures were captured during SEEG, the pre-SEEG seizure-onset hypothesis was focal-local in 51 patients (47.2%), focal-regional in 18 (16.7%), focal-multiregional in 31 (28.7%), and unknown in eight (7.4%). No patient had a pre-SEEG hypothesis of generalized epilepsy; at our institution, patients with idiopathic generalized epilepsy are offered CM-RNS without thalamic SEEG. Following SEEG, the cortical seizure-onset hypothesis was revised in 59 of 108 patients (54.6%; partially revised in 45.4% and fully revised in 9.3%) (Figure S1).

Of the 51 patients with a focal-local hypothesis, 22 showed no early thalamic involvement, and 19 of these underwent resection or laser interstitial thermal therapy (LITT). The remaining three, in whom the seizure-onset zone (SOZ) involved eloquent cortex or was too broad, received neuromodulation. Eighteen patients showed unilateral early thalamic involvement, of whom eight were not candidates for an ablative procedure and received corticocortical RNS (n = 2), thalamocortical RNS (n = 4), or thalamic RNS (n = 2). Of the 11 patients with bilateral early thalamic involvement, nine received thalamic or thalamocortical RNS; the exceptions were a patient who underwent resection of a parietal SOZ that propagated rapidly to both thalami and a patient with bitemporal epilepsy who received bihippocampal RNS. Of the 17 patients in this group whose treatment plan included thalamic or thalamocortical RNS, four had a revised target (Figure 4C).

**Figure 3.**
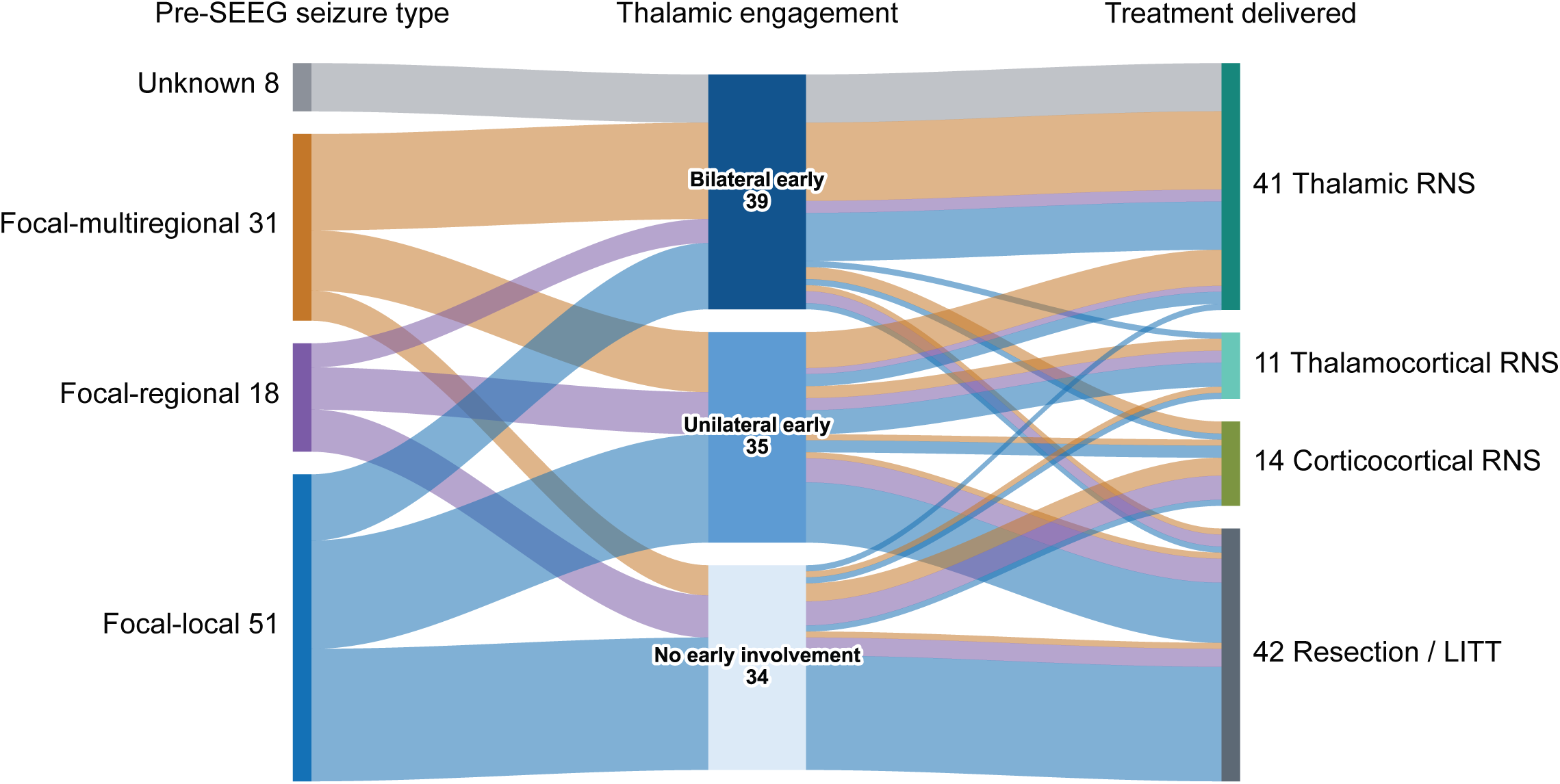
Thalamic network laterality and ictal timing inform the RNS strategy. Three-stage alluvial diagram for all patients who had a spontaneous seizure during the SEEG EMU stay (n = 108 patients; for the five patients still planning therapeutic surgery, the planned treatment is shown). Ribbons retain the color of their pre-SEEG hypothesized seizure type across both transitions, so a single seizure-type group can be traced to the therapy it ultimately received; ribbon width is proportional to the number of patients, and node labels give the count in each category. *Left (pre-SEEG seizure type)*: focal-local 51, focal-multiregional 31, focal-regional 18, unknown 8. *Middle (thalamic engagement on SEEG)*: early bilateral 39, early unilateral 35, no early thalamic involvement 34. *Right (treatment delivered)*: resection/LITT 42, thalamic RNS 41, thalamocortical RNS 11, corticocortical RNS 14.

**Figure 4.**
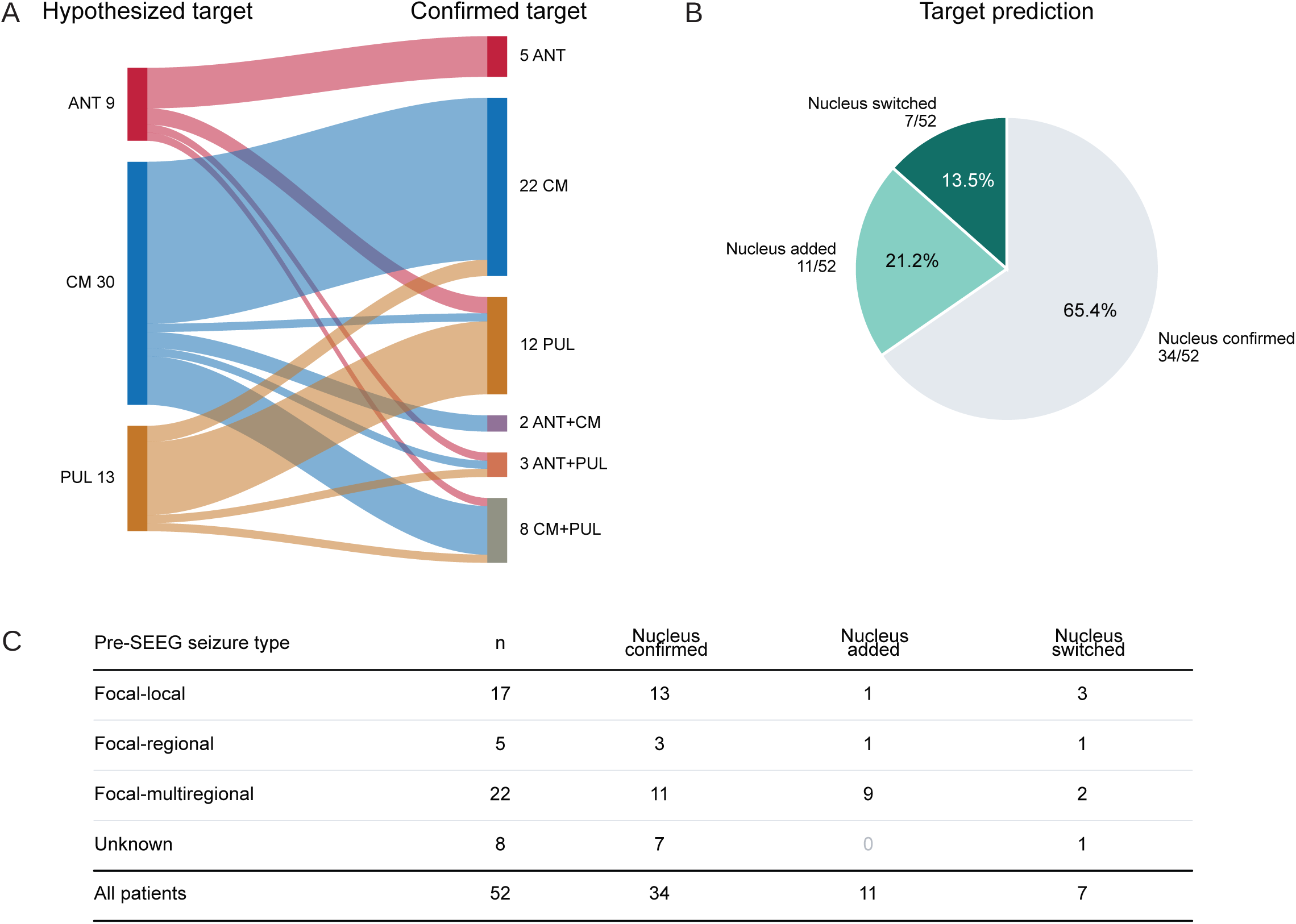
Thalamic SEEG informs the RNS target. Analysis is restricted to the 52 patients whose treatment plan included thalamic or thalamocortical RNS (48 patients received treatment, and four patients are awaiting surgery). **(A)** Pre-SEEG hypothesized thalamic target versus the confirmed thalamic target, as a two-stage alluvial diagram. Ribbons retain the color of the hypothesized nucleus (ANT red, CM blue, PUL amber; combination targets are the blend of their constituent hues), and node labels give the number of patients. **(B)** Thalamic neuromodulation target prediction accuracy. The pre-SEEG hypothesized thalamic target was confirmed in 34 of 52 patients (65.4%), an additional thalamic nucleus was added to the hypothesized target in 11 (21.2%), and the target was switched to a different nucleus in seven (13.5%). **(C)** Revision of thalamic neuromodulation targets through thalamic SEEG for each pre-SEEG hypothesized seizure type.

Among the 18 patients with a focal-regional hypothesis, every patient with early thalamic involvement in whom ablation was not feasible received thalamic or thalamocortical RNS (n = 5), whereas every patient without early thalamic involvement and without an ablative target received corticocortical RNS (n = 4). The remaining nine patients underwent an ablative procedure. Of the five patients who received thalamic or thalamocortical RNS, two had a revised target (Figure 4C).

Among the 31 patients with a focal-multiregional hypothesis, 13 of the 16 with bilateral early thalamic involvement received thalamic RNS, two of the 16 received bihippocampal RNS, and one patient underwent resection of the dominant focus with staged thalamic RNS planned. Of the 10 patients with unilateral early thalamic involvement, eight received thalamic or thalamocortical RNS, one received bihippocampal RNS, and one underwent resection. One of the five patients without early thalamic involvement received thalamocortical RNS because the thalamic signal, although delayed relative to cortical onset, preceded clinical seizure manifestation. The thalamic target was revised in 11 of 22 patients (50.0%) in this group, the highest revision rate of any seizure type (Figure 4C); in 9 of these 11, an additional nucleus was added to the hypothesized target after thalamic SEEG demonstrated involvement of different nuclei across seizure types.

All eight patients with an unknown seizure type showed bilateral early thalamic involvement and underwent thalamic RNS. The planned target was unchanged in seven; in the remaining patient, who had a frontotemporal onset pattern, the target was revised from CM to combined ANT+PUL.

Across the cohort, the thalamic target was switched in seven of 52 patients (13.5%) and augmented with an additional nucleus in eleven (21.2%; Figure 4). We then asked whether the thalamic target changed only when the cortical seizure-onset hypothesis also changed. It did not. Among the 15 patients whose cortical hypothesis was unchanged after SEEG, the thalamic target was revised in five (Figure 5A).

**Figure 5.**
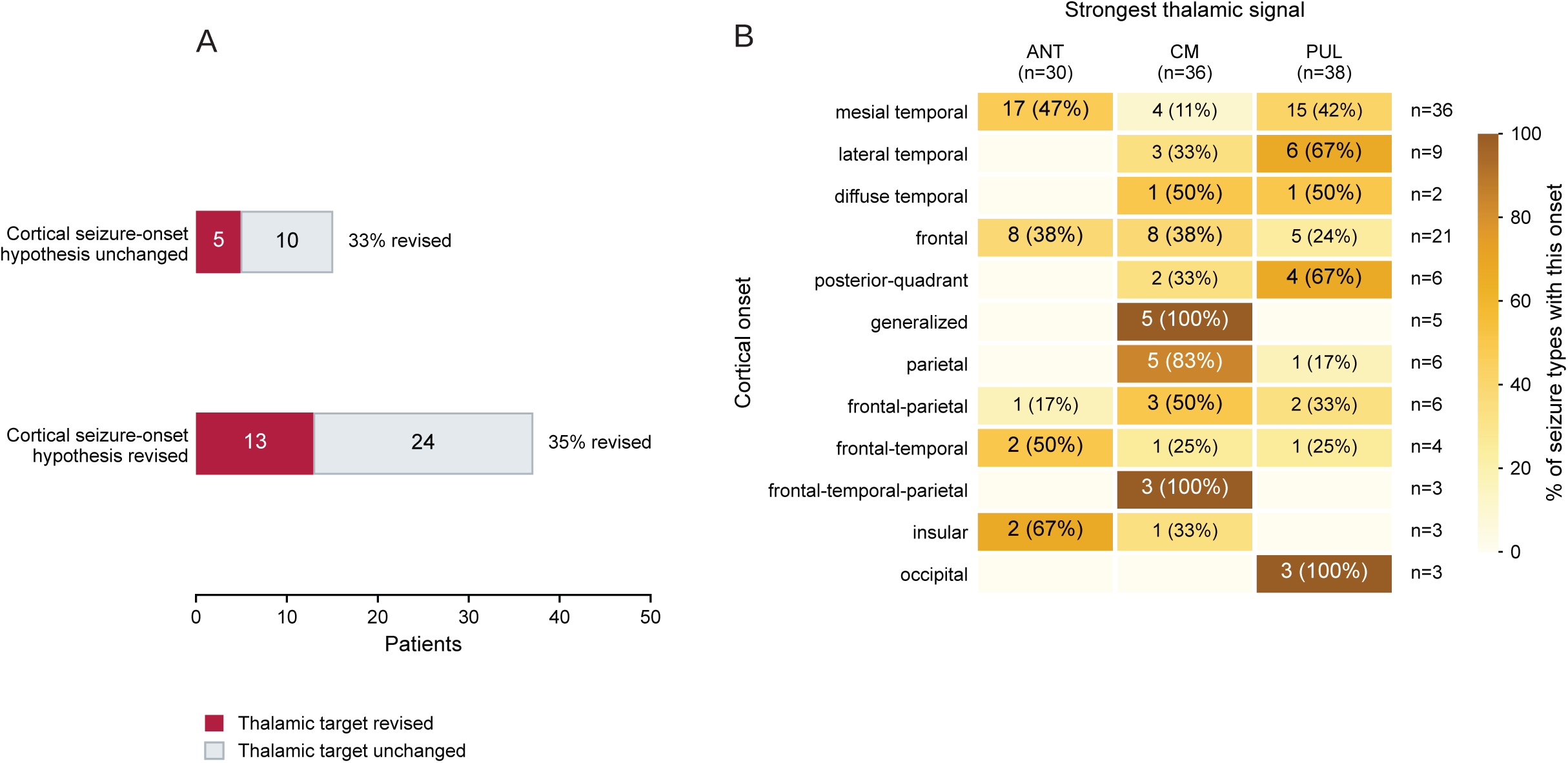
Cortical seizure-onset does not always predict the leading thalamic nucleus. **(A)** Thalamic target revision according to whether the cortical seizure-onset hypothesis was revised, in 52 patients planned for thalamic or thalamocortical RNS. Bars are split by whether the cortical seizure-onset hypothesis was unchanged (n = 15) or revised (n = 37) after SEEG; the red segment of each bar is the number whose thalamic target was revised. The thalamic target was revised in five of 15 patients (33.3%) whose cortical hypothesis was unchanged and in 13 of 37 (35.1%) whose cortical hypothesis was revised, indicating that thalamic target revision was not simply a downstream consequence of cortical seizure-onset hypothesis revision. **(B)** Heatmap of cortical seizure-onset zone versus the thalamic nucleus with the strongest ictal signal. The unit of analysis is the seizure type rather than the patient (104 seizure types from 75 patients; a single patient can have multiple seizure types), and the analysis is restricted to patients in whom all thalamic SEEG contacts lay within the intended nucleus (accurate targeting) on lead reconstruction and who had thalamic involvement during seizures. Each cell gives the number of seizure types with that cortical onset whose strongest thalamic signal was in that nucleus, with the percentage of that onset’s seizure types in parentheses; cell shading follows this row percentage. Row totals are given on the right and nucleus totals at the top. The posterior-quadrant category includes temporal-parietal, temporal-occipital, and parietal-occipital seizure onset. Most cortical seizure-onset zones were associated with more than one thalamic nucleus.

We then compared the cortical seizure-onset location with the thalamic nucleus showing the strongest ictal signal, restricting the analysis to patients in whom every thalamic lead lay within its intended nucleus (Figure 5B). The leading nucleus varied within each onset location, and no cortical region mapped consistently onto a single nucleus, except for generalized onset and diffuse frontal-temporal-parietal onset, which were associated with CM in all eight instances, and occipital onset, which was associated with PUL in all three instances. Cortical seizure-onset information alone therefore does not reliably predict the optimal thalamic target for neuromodulation.

### Early thalamic signal is associated with better outcome after neuromodulation

Of the 103 patients who underwent therapeutic surgery after SEEG, 86 had seizure outcome assessment more than 6 months after the most recent procedure (excluding one patient whose RNS was removed because of infection). The mean follow-up duration was 24.1 ± 15.8 months. Seventeen patients underwent more than one procedure (seven planned, 10 for incomplete seizure control). After all interventions, mean primary seizure reduction was 81.3 ± 28.8%, with freedom from primary seizure type achieved in 48.8% of patients (Figure 6A–C). Mean primary seizure reduction was 90.9 ± 20.6% after resection or LITT (seizure freedom 61.8%) and 75.0 ± 31.7% after RNS, including RNS after an ablative procedure (seizure freedom 40.4%). The 12 with outcome data who received RNS as a staged or second procedure had a mean primary seizure reduction of 67.3 ± 36.9% (seizure freedom 33.3%).

**Figure 6.**
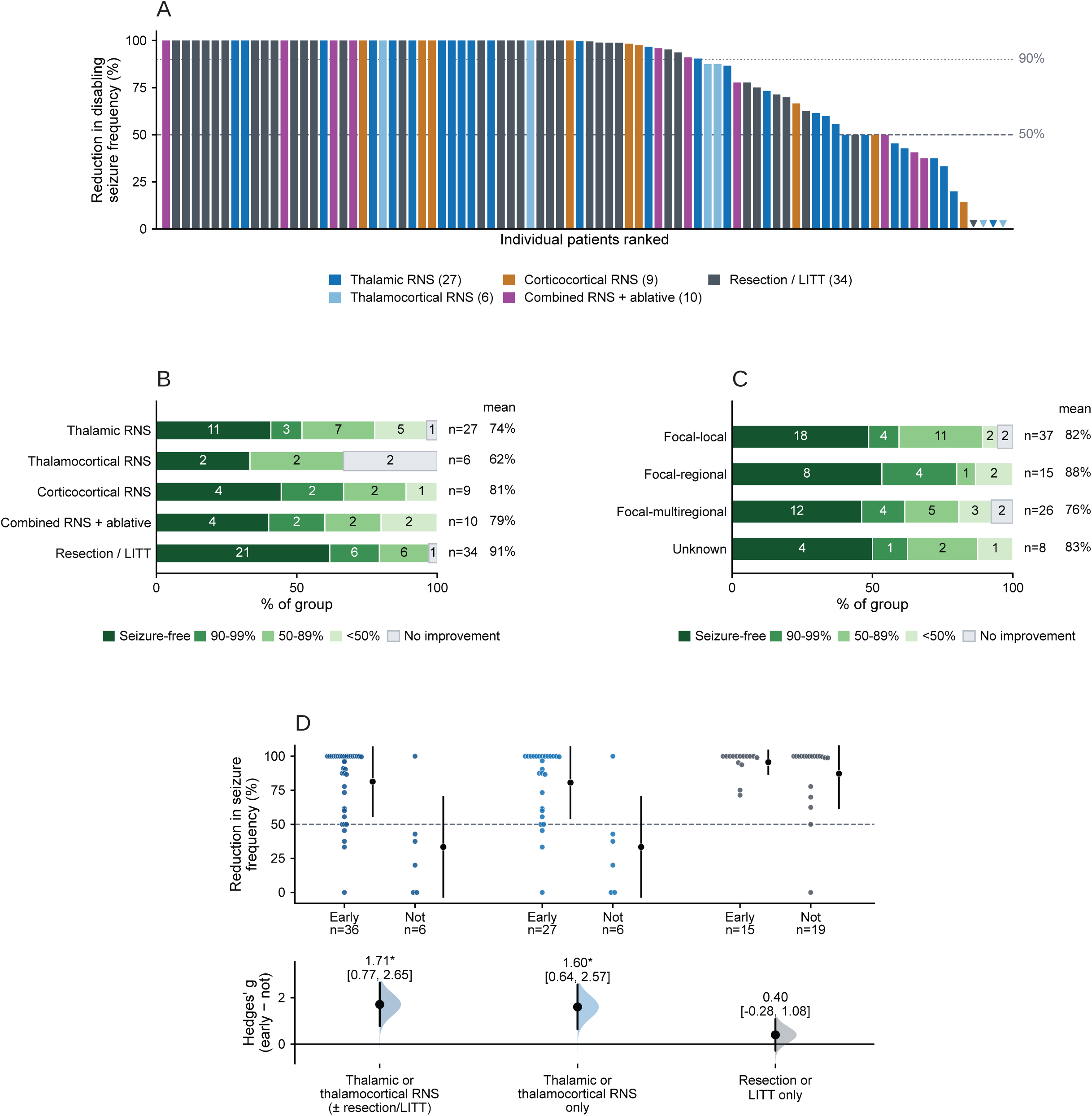
Early thalamic engagement is associated with response to neuromodulation but not ablation. All panels use 86 patients with a recorded percentage reduction in seizure frequency of the primary seizure type after at least 6 months of follow-up after their last therapeutic surgery (mean follow-up 24.1 ± 15.8 months). **(A)** Waterfall plot of the percentage reduction in seizure frequency for every patient, ranked from greatest to least reduction and colored by the treatment received (thalamic RNS, thalamocortical RNS, corticocortical RNS, combined RNS + ablative, resection/LITT). **(B)** Primary seizure frequency reduction by treatment group: seizure-free, 90–99%, 50–89%, <50% reduction and no improvement (seizure frequency unchanged or worse). Thalamic RNS n = 27 (11 seizure-free; mean reduction 74%), thalamocortical RNS n = 6 (2; 62%), corticocortical RNS n = 9 (4; 81%), combined RNS + ablative n = 10 (4; 79%), resection/LITT n = 34 (21; 91%). **(C)** Primary seizure frequency reduction by pre-SEEG hypothesis type: focal-local n = 37 (18 seizure-free; mean 82%), focal-regional n = 15 (8; 88%), focal-multiregional n = 26 (12; 76%), unknown n = 8 (4; 83%). **(D)** Primary seizure frequency reduction by thalamic ictal signal timing (early vs not early), within three treatment groups. Individual patients are shown with the group mean ± SD (upper plot); the lower plot gives the standardized mean difference (Hedges’ g, early − not early) with its 95% CI, and an asterisk marks a CI that excludes zero. Among patients treated with thalamic or thalamocortical RNS, including those who also underwent resection or LITT, an early thalamic signal was associated with a greater reduction in seizure frequency (early n = 36 vs not early n = 6; Hedges’ g 1.71, 95% CI 0.77–2.65), and the association persisted when the analysis was restricted to thalamic or thalamocortical RNS alone (early n = 27 vs not early n = 6; g = 1.60, 95% CI 0.64–2.57), whereas no such association was seen among patients treated with resection or LITT alone (early n = 15 vs not early n = 19; g = 0.40, 95% CI −0.28 to 1.08).

We next investigated whether early thalamic ictal signals during SEEG were associated with seizure outcomes following RNS (Figure 6D). Among patients who received thalamic or thalamocortical RNS, those with early thalamic signals had better seizure outcomes than those without early thalamic signals (early n = 36, not early n = 6; Hedges’ g 1.71, 95% CI 0.77–2.65). The association persisted when patients who received both RNS and an ablative procedure were excluded (Figure 6D, early n = 27, not early n = 6; g 1.60, 95% CI 0.64–2.57). In contrast, seizure outcomes did not differ between patients with and without early thalamic signals among those who underwent ablative treatment (early n = 15, not early n = 19; g 0.40, 95% CI −0.28 to 1.08).

## DISCUSSION

In this retrospective series of 117 patients undergoing thalamic SEEG at Massachusetts General Hospital, we evaluated the safety, targeting accuracy, clinical utility, and seizure outcomes associated with implantation of the ANT, CM, and PUL thalamic nuclei as part of phase 2 epilepsy evaluation.

We found that thalamic SEEG implantation has a safety profile comparable to non-thalamic SEEG. No hemorrhage was symptomatic or required surgical intervention, and none was associated with a thalamic lead. The hemorrhage rate is in line with the 0.5% per lead reported for SEEG overall^20^ as well as a published scoping review of 243 thalamic SEEG cases across 26 centers, which reported no permanent neurological deficits attributable to thalamic trajectories and asymptomatic hemorrhages in five cases.^9^ Our findings are also consistent with the foundational thalamic SEEG series of Talairach and Bancaud, in which 144 patients underwent thalamic implantation without lasting morbidity.^21^

Our findings also provide empirical support for the concept that thalamic investigation can serve as “hemispheric surveillance that leverages the condensed thalamic map of the widespread cortex spanned by the circuit.” ^22^ Our results suggest that sampling the contralateral thalamus may provide an efficient means of assessing possible contralateral network involvement when the primary seizure-onset hypothesis is unilateral but there is concern for an additional contralateral seizure onset. Bitemporal epilepsy may not recruit the thalamus to the extent that other bilateral seizure networks do, and the sensitivity of early bilateral thalamic signals for detecting bilateral seizure network rose to 92.6% when bitemporal cases were excluded.

Our results further demonstrate that thalamic SEEG can inform both neuromodulation strategy and thalamic target selection. This follows from the architecture of RNS: because each lead is programmed independently for detection and stimulation, the configuration most likely to disrupt a patient’s seizure organization depends on which contacts see the seizure first.^13^ Thalamic sampling therefore enters the SEEG plan as the means of testing whether a thalamic node can serve as detector, stimulator, or both, and whether the network is unilateral or bilateral. The primary question addressed by thalamic SEEG is the timing of thalamic recruitment relative to cortical onset, and this series is an empirical test of the framework in which that timing selects the RNS configuration.^10^ When the thalamus was recruited early relative to cortical seizure onset and there was no localized SOZ amenable to resection or LITT, thalamic or thalamocortical RNS was selected. In contrast, corticocortical RNS was selected when early thalamic involvement was absent or late. Exceptions occurred, mostly in focal-multiregional epilepsy, when delayed thalamic recruitment (>1 second) still preceded clinical seizure onset. These exceptions indicate that the one-second threshold is a working definition rather than a validated cutoff; the lag that best predicts response to each configuration remains to be determined empirically.^10^ In all cases, treatment decisions were based on the integrated clinical and electrophysiological evaluation rather than thalamic recordings alone.

Thalamic SEEG also frequently altered the hypothesized neuromodulation target. The thalamic target was revised in 34.6% of cases overall, with the highest rate of revision occurring among patients with focal-multiregional seizure type (50.0%), but target revisions also occurred in patients with focal-local seizure type (23.5%). Target revision did not require revision of the cortical seizure-onset hypothesis; among patients whose cortical hypothesis was unchanged, the thalamic target was revised in 33.3%. Furthermore, our analysis of cortical seizure-onset locations and the thalamic nuclei demonstrating the strongest ictal signals showed that most cortical onset locations lacked a one-to-one relationship with thalamic recruitment. While knowledge of thalamocortical connectivity can help identify plausible thalamic targets for neuromodulation,^23^ our findings suggest that direct thalamic recording may provide additional patient-specific information that cannot be reliably inferred from cortical SEEG alone, particularly in complex multiregional seizure networks. The MORE experience^11^ points to the same conclusion: effectiveness fell when stimulation was applied without regard to whether the Papez circuit participated in each patient’s seizure organization.

Our outcome analysis also identified an association between early thalamic recruitment and seizure response to neuromodulation. Among patients treated with thalamic or thalamocortical RNS, those with early thalamic ictal signals had better seizure outcomes than those without early thalamic signals (Figure 6D). Although this analysis is retrospective and does not establish a causal relationship, the finding supports our practice of offering thalamic neuromodulation when early thalamic involvement is demonstrated, and it agrees with the report that stimulation of hodologically matched thalamic nuclei produced better seizure outcomes.^23^ Their approach matched nuclei to seizure-onset zones through structural and functional connectivity, which they termed thalamocortical hodology (hodology, the study of pathways). Our method extends this concept to ictal hodology: anatomical connectivity nominates candidate thalamic nuclei, and the timing of their ictal recruitment identifies the nucleus that participates most in a patient’s seizure network. There was no corresponding difference among patients treated with resection or LITT, in contrast to one study that linked estimated early thalamic involvement to poor outcomes after ablative treatment.^24^

Our thalamic sampling strategy evolved over the study period. This was one component of a broader shift at our center from focus-directed to network-directed SEEG, over which the proportion of evaluated patients offered no surgical option fell from 22% to 3%.^25^ Each implantation was planned on a patient-by-patient basis to resolve a specific diagnostic question or to inform a candidate RNS strategy. Thalamic sampling undertaken on this basis falls within clinical practice rather than research, and the safety and decision-making data reported here support its incorporation into routine evaluation whenever the clinical hypothesis demands it.^10^

Several limitations of this study warrant consideration. First, this was a single-center retrospective study, which may limit the generalizability of our findings. Second, postoperative outcome data were available for only 86 of 103 patients who underwent therapeutic intervention, with a mean follow-up of 24.1 ± 15.8 months. This duration may be insufficient to characterize the long-term efficacy of RNS, whose benefit may continue to evolve over several years^26^, or late seizure recurrence after ablative treatment. Third, visual review of SEEG was not blinded to patient information. Finally, because the study was not designed as a prospective comparison of different SEEG targeting strategies or neuromodulation approaches, the observed associations between thalamic activity and treatment selection or outcome should be interpreted as hypothesis-generating.

In conclusion, this study provides the largest modern single-center series of thalamic SEEG and demonstrates that hypothesis-driven thalamic sampling is safe and informs treatment decisions. These findings support its use in the evaluation of patients with drug-resistant epilepsy whenever network laterality or a thalamic neuromodulation target is in question. Because both RNS and ANT-DBS reduce the incidence of sudden unexpected death in epilepsy by about two thirds^3,4^, accurate identification of the optimal thalamic node for each patient has implications that extend beyond seizure reduction alone.

## Acknowledgements

This work was supported by R61NS126776 (RMR) and UE5NS065743 (SL).

## Conflicts of Interest

R.M.R. is a consultant for NeuroPace, Inc. and Zimmer Biomet, Inc. The remaining authors have no conflicts of interest.

## Ethical Publication Statement

We confirm that we have read the Journal’s position on issues involved in ethical publication and affirm that this report is consistent with those guidelines.

## Data Availability Statement

De-identified data are available from the corresponding author on reasonable request.

---

**Figure S1.**
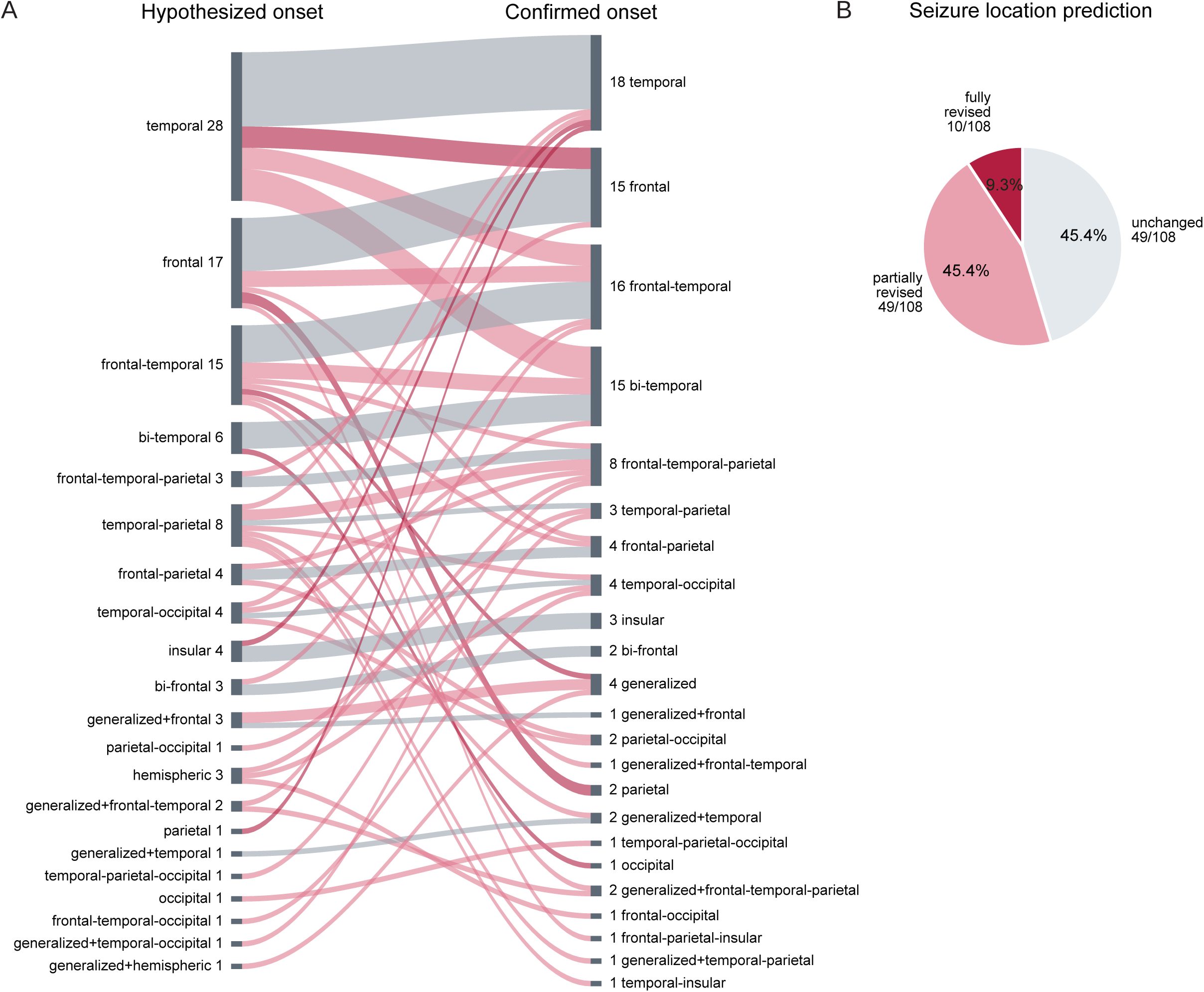
Revision of the seizure-onset location hypothesis after SEEG. **(A)** Pre-SEEG hypothesized cortical seizure-onset location versus the seizure-onset location confirmed after SEEG, as a two-stage alluvial diagram. Ribbons are grey where the pre-SEEG hypothesized location was confirmed by SEEG, light red where it was partially revised (the pre- and post-SEEG locations differ but share at least one region) and dark red where it was fully revised (no region in common). Node labels give the number of patients (n = 108). **(B)** Seizure-onset location prediction accuracy. The pre-SEEG hypothesized location was retained unchanged in 49 of 108 patients (45.4%), partially revised in 49 (45.4%) and fully revised in 10 (9.3%).

**Table S1:**

| **Characteristic** |  | **All patients (n=117)** |
|---|---|---|
| Patients, n |  | 117 |
| Age at SEEG (years) | Mean ± SD | 27.8 ± 14.8 |
|  | Median (range) | 24.0 (1–74) |
| Epilepsy duration (years) | Mean ± SD | 13.2 ± 9.3 |
|  | Median (range) | 12.0 (0–48) |
| Etiology, n (%) |  |  |
|  | MRI-lesional | 49 (41.9%) |
|  | not known | 48 (41.0%) |
|  | post-traumatic | 9 (7.7%) |
|  | post-infectious | 5 (4.3%) |
|  | genetic | 4 (3.4%) |
|  | post-hemorrhage | 2 (1.7%) |
| Seizure onset location, n (%) [n=108] |  |  |
|  | temporal | 33 (30.6%) |
|  | frontal | 17 (15.7%) |
|  | frontal-temporal | 16 (14.8%) |
|  | frontal-temporal-parietal | 8 (7.4%) |
|  | generalized with focal features | 7 (6.5%) |
|  | generalized | 4 (3.7%) |
|  | frontal-parietal | 4 (3.7%) |
|  | temporal-occipital | 4 (3.7%) |
|  | temporal-parietal | 3 (2.8%) |
|  | insular | 3 (2.8%) |
|  | parietal | 2 (1.9%) |
|  | parietal-occipital | 2 (1.9%) |
|  | occipital | 1 (0.9%) |
|  | temporal-insular | 1 (0.9%) |
|  | temporal-parietal-occipital | 1 (0.9%) |
|  | frontal-parietal-insular | 1 (0.9%) |
|  | frontal-occipital | 1 (0.9%) |
| Treatment type, n (%) [n=108] |  |  |
|  | Thalamic RNS | 31 (28.7%) |
|  | Thalamocortical RNS | 8 (7.4%) |
|  | Corticocortical RNS | 13 (12.0%) |
|  | Combined RNS + ablative | 10 (9.3%) |
|  | Resection / LITT | 41 (38.0%) |
|  | No therapeutic surgery yet | 5 (4.6%) |
| Excluded from analysis, n (%) |  |  |
|  | No (spontaneous seizure captured) | 108 (92.3%) |
|  | Yes (no spontaneous seizure captured) | 9 (7.7%) |

## Notes

### Author Declarations

The study was approved by the Mass General Brigham Institutional Review Board, which waived the requirement for informed consent for this retrospective review.

